# Pregnancy loss and postpartum mortality in a sub-Saharan African setting

**DOI:** 10.1101/2021.03.18.21253872

**Authors:** Yoshan Moodley, Hallie Eilerts, Kobus Herbst, Frank Tanser

**Author notes:** **Correspondence:** Yoshan Moodley (PhD), Africa Health Research Institute, Private Bag X7, Congella 4013, South Africa. **Funding:** The corresponding author was supported with a postdoctoral fellowship under a National Institute of Health (NIH) grant (R01 HD084233). The Africa Health Research Institute’s (AHRI’s) Demographic Surveillance Information System and Population Intervention Programme is funded by the Wellcome Trust (201433/Z/16/Z), and the South African Population Research Infrastructure Network (funded by the South African Department of Science and Technology, hosted by the South African Medical Research Council). The funders had no role in designing this study; analysis or interpretation of the data; writing this manuscript; or the decision to submit the article for publication. **PRECIS** Pregnancy loss is associated with increased postpartum mortality in sub-Saharan African women, highlighting the need for postpartum medical care and psychological support in this group.

## Abstract

**Objective:** Studies from industrialized countries report a harmful association between pregnancy loss and postpartum mortality. However, the nature of this relationship has not been established in resource limited sub-Saharan Africa. Given the potential implications of this knowledge for risk stratification and subsequent maternal health in sub-Saharan Africa, our study sought to use data from one of the continent’s largest and longest running population-based cohorts to investigate the relationship between pregnancy loss and postpartum mortality.

**Methods:** We conducted a population-based cohort study involving 25048 postpartum women from KwaZulu-Natal, South Africa. The study data was collected during biannual/triannual general household surveys, which also incorporated a pregnancy questionnaire for women who reported a pregnancy between survey waves. Pregnancy loss was defined as a pregnancy which ended in abortion, miscarriage, or stillbirth. Postpartum mortality was defined as the death of a woman, from any cause, within one year after the date that her pregnancy ended. We used a quasi-poisson regression model, adjusted for maternal age and other setting-specific predictors of postpartum mortality, to investigate the relationship between pregnancy loss and postpartum mortality.

**Results:** The incidence of postpartum mortality was three times higher in women who experienced pregnancy loss (Adjusted Incidence Rate Ratio: 3.23, 95% Confidence Interval: 2.13-4.71), when compared with women who had live births.

**Conclusion:** Our results reveal a clear association between pregnancy loss and increased postpartum mortality in a typical rural sub-Saharan African setting. Women who have recently experienced pregnancy loss should be targeted with a package of risk reduction interventions, including additional opportunities for medical care or psychosocial support.

## INTRODUCTION

The current population of sub-Saharan Africa, which was estimated at 1.07 billion people in 2019, is expected to double by the year 2050.^1^ On average, each sub-Saharan African woman will have between 4 and 5 children during her lifetime.^1^ Pregnancy and childbirth is not without risk,^2, 3^ and postpartum mortality will continue to be of public health relevance in this resource-constrained region for many years to come.

Maternal mortality rates in sub-Saharan African countries, such as South Africa, are far higher when compared with industrialized countries.^3^ Although the rollout of HIV antiretroviral therapy in rural KwaZulu-Natal, South Africa has contributed to a reduction in the number of maternal deaths in this setting, this number is still excessively high and requires further attention.^4-6^ Almost 60% of maternal mortality in South Africa is preventable,^5^ highlighting the potential benefits of using risk stratification methods based on modifiable or non-modifiable risk factors to identify high-risk women, who could then be efficiently targeted with appropriate risk reduction interventions. ^7^

A recent study from rural KwaZulu-Natal, South Africa by Tlou and colleagues found that HIV serostatus, level of education, and prior parity are strong predictors of maternal mortality.^6^ Although Tlou et al.,^6^ did not include pregnancy loss (which encompasses abortion, miscarriage, and stillbirth) as a variable of interest in their analysis, the international literature reports a significantly higher risk of postpartum mortality in women who have recently experienced pregnancy loss.^8, 9^ If the findings from the international literature also hold true for rural KwaZulu-Natal, South Africa then it will have dire consequences for maternal survival in this setting, as existing rates of pregnancy loss in South Africa are alarmingly high.^10, 11^ However, there have not been any published studies thus far which have sought to provide a clear understanding of the relationship between pregnancy loss and postpartum mortality in resource limited sub-Saharan Africa. Given the implications of this knowledge for risk stratification and subsequent maternal health in sub-Saharan Africa, our study sought to use data from one of Africa’s largest and longest running population-based cohorts to investigate the relationship between pregnancy loss and postpartum mortality.

## METHODS

### Study design and setting

We conducted a population-based, prospective cohort study of postpartum women from a community in rural KwaZulu-Natal, South Africa. The community has been part of the Africa Health Research Institute (AHRI) health surveillance platform for almost 20 years and has been described in detail elsewhere.^12^

### Data sources

Data for AHRI’s ongoing health surveillance platform is collected through ongoing population-based general household surveys.^12^ These general household surveys were initially conducted on a biannual basis between 2000 and 2011, but have been conducted triannually since 2012.^12, 13^ Data for the AHRI general household surveys is collected by trained research assistants. The data is then entered onto electronic databases, managed by dedicated data management teams. Quality control of the data is implemented on a regular basis. Response rates for the AHRI general household surveys are usually >98%. The general household surveys include a pregnancy questionnaire for women who report pregnancies between survey waves.^13^ The pregnancy questionnaire collects data on maternal characteristics, such as age, date of last menses, and prior maternal history; and pregnancy characteristics, such as the date that the pregnancy ended and the outcome of the pregnancy.^13^ Data on pregnancy outcomes are predominantly collected using a single item on the pregnancy questionnaire – “Was the baby born alive, born dead (i.e. stillbirth), or did the pregnancy end in a miscarriage or abortion?”. Only one of the aforementioned pregnancy outcomes can be selected for each completed pregnancy questionnaire. Maternal deaths and dates on which these deaths occurred are recorded during participant follow-up at subsequent waves of the general household survey.^12^ Information on the death of a general household survey participant is usually derived from other members of the household where the deceased participant resided. Data for other known predictors of maternal mortality in rural KwaZulu-Natal, such as HIV serostatus and the woman’s level of education, are collected as part of other questionnaires comprising the general household survey.^12^

### Measures

Pregnancy loss was defined as any pregnancy which did not result in a live birth. This included pregnancies which ended in an abortion, a miscarriage, or a stillbirth. Pregnancy loss was predominantly established from the responses provided to the question “Was the baby born alive, born dead, or did the pregnancy end in a miscarriage or abortion?” on the pregnancy questionnaire. As further clarification of the definitions for the individual pregnancy outcomes used in this study, live births were defined as infants born alive; stillbirths were defined as infants that did not demonstrate any signs of life at the time of birth; abortions were defined as any elective termination of pregnancy (the vast majority of abortions in our setting are elective abortions for unwanted pregnancies rather than therapeutic abortions for pregnancy complications which threaten the life of the mother); and miscarriages were defined as pregnancies of <28 weeks duration that were not marked as abortions or stillbirths on the pregnancy questionnaire.

The study outcome, postpartum mortality, was defined as the death of a woman from any cause, within one year after the date that her pregnancy ended. This was established from subsequent waves of the general household survey. For a woman who had suffered postpartum mortality, the follow-up time (in woman years – wy) was calculated as the difference in time between the date of her death and the date that her pregnancy ended. Women who did not suffer postpartum mortality were censored at one year after the date that their pregnancy ended.

### Eligibility criteria

We included records from AHRI population-based surveys for all women aged ≥16 years old who reported singleton pregnancies between 1 January 2000 and 31 December 2017. The derivation of the study sample is outlined in Appendix 1.

Briefly, there were 83720 women aged ≥16 years old residing in the AHRI study area between 2000 and 2017. Of these 83720 women, 82907 agreed to participate in the general household survey and provide pregnancy information during subsequent waves of the survey (equating to a response rate of 99%). The study sample was comprised of 25048 women after excluding women who did not report pregnancies during the study period, women who had twins, and subsequent pregnancies for the same woman (i.e. only the first recorded pregnancy during the study period was used for each woman).

### Data analysis

We used R version 3.6.2 (R Foundation for Statistical Computing, Vienna, Austria) to perform the data analysis. We used descriptive statistical methods to summarize the characteristics of our study sample and present these summary statistics as frequencies and percentages. The continuous variable “Maternal age” was categorized into 3 groups: 16-25 years old, 26 to 35 years old, and >35 years old. We performed crude and adjusted quasi-poisson regression analyses to investigate the relationship between pregnancy loss and postpartum mortality. The adjusted quasi-poisson regression analysis was controlled for maternal age and previously identified predictors of maternal mortality in rural KwaZulu-Natal, namely HIV serostatus, level of education, and parity.^6^ We present the results of the quasi-poisson regression analyses as incidence rate ratios (IRRs) with 95% confidence intervals (95% CI).

### Ethical approval

AHRI’s ongoing population-based health surveillance platform has received approval from the Biomedical Research Ethics Committee at the University of KwaZulu-Natal, South Africa (Protocol BE290/16). Written informed consent is required prior to completing the general household surveys.

## RESULTS

The characteristics of the study sample, comprised of 25048 women, are shown in Table 1. The majority of women were aged between 16 and 25 years old (n=17517, 69.9%). Just over half of all women had completed primary school (n=13936, 55.6%). An HIV-positive serostatus was recorded for 1595 women (6.4%). Most women had a prior history of pregnancy (n=23659, 94.5%). A total of 941 women (3.8%) experienced pregnancy loss, while the remaining 24107 women (96.2%) had live births. Of the 941 lost pregnancies, 484 (51.4%) were abortions, 271 were stillbirths (28.8%), and 186 were miscarriages (19.8%). The total follow-up time for the cohort was 24944.94 wy. We were able to establish postpartum mortality status for 100% of women in the study sample (i.e. there were no women in our study with missing postpartum mortality information). A total of 178 women (0.7% of the study sample) died, and the crude incidence of postpartum mortality was estimated at 7.14 (95% CI: 6.15-8.28) per 1000wy.

**Table 1.** Description of the study sample (N=25048)

| Characteristic | Summary statistic |
| --- | --- |
| *Age categories, n (% of N)* |  |
| 16-25 years old | 17517 (69.9) |
| 26-35 years old | 5977 (23.9) |
| >35 years old | 1154 (6.2) |
| *Completed Primary School, n (% of N)* |  |
| No | 11112 (44.4) |
| Yes | 13936 (55.6) |
| *HIV-positive, n (% of N)* |  |
| No | 23453 (93.6) |
| Yes | 1595 (6.4) |
| *\*Prior parity, n (% of N)* |  |
| No | 1389 (5.5) |
| Yes | 23659 (94.5) |
| *Pregnancy loss, n (% of N)* |  |
| No | 24107 (96.2) |
| Yes | 941 (3.8) |

\*Does not include the index pregnancy used in this study.

The rate of postpartum mortality, stratified by various maternal characteristics, is shown in Table 2. The crude rate of postpartum mortality was higher in women who experienced pregnancy loss (Crude rate: 21.57 [95% CI: 13.58-33.74] per 1000wy) versus women who had live births (Crude rate: 6.58 [95% CI: 5.61-7.71] per 1000wy). A quasi-poisson analysis, adjusted for established predictors of maternal mortality in our setting (maternal age, level of education, parity, and HIV serostatus),^6^ found that postpartum mortality was three-times higher in women who experienced pregnancy loss (Adjusted IRR: 3.23, 95% CI: 2.13-4.71).

**Table 2.**
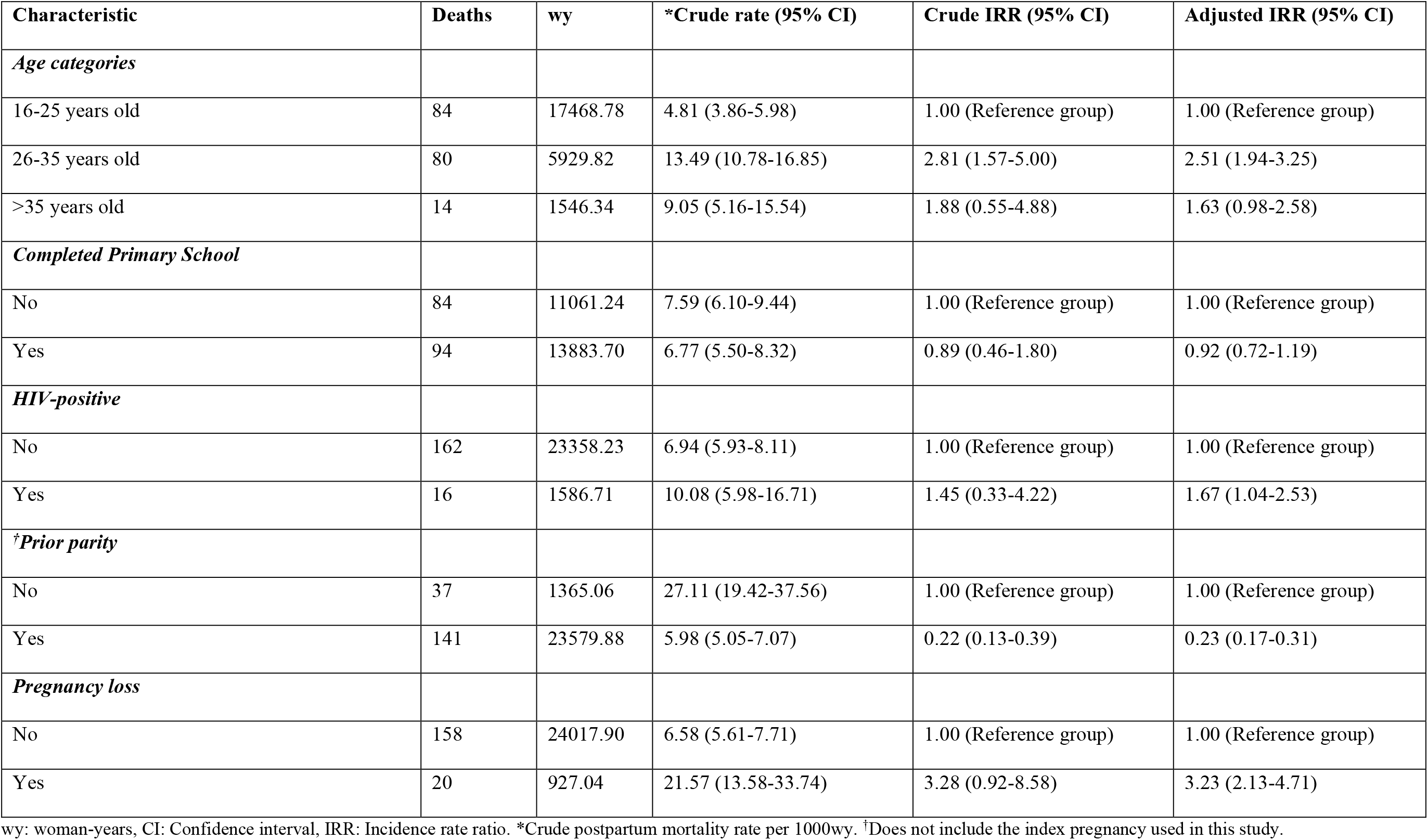
Rate of postpartum mortality in the study sample (N=25048), stratified by maternal characteristics

The adjusted quasi-poisson analysis also revealed increased postpartum mortality in women aged 26-35 years old (Adjusted IRR: 2.51, 95% CI: 1.94-3.25) and HIV-positive women (Adjusted IRR: 1.67, 95% CI: 1.04-2.53). Postpartum mortality was lower in women who reported prior parity (Adjusted IRR: 0.23, 95% CI: 0.17-0.31).

## DISCUSSION

Our analysis of data from over 25000 postpartum women who participated in one of Africa’s longest running and largest population-based cohorts suggests a three-fold higher incidence of postpartum mortality in women who experienced pregnancy loss when compared with women who had live births. A recent systematic review and meta-analysis by Reardon and Thorp reported that the risk of postpartum mortality was two-fold higher (pooled odds ratio: 2.37, 95% CI: 2.04-2.76) in northern European women who suffered a pregnancy loss when compared with women who had live births.^9^ Thus, our study confirms the harmful association between pregnancy loss and postpartum mortality reported in Reardon and Thorp’s meta-analysis,^9^ highlighting the importance of this pregnancy outcome on subsequent maternal survival in both industrialized and developing countries.

Maternal mortality is multifactorial.^6^ Therefore, pregnancy loss is one of several factors which contribute to postpartum mortality in our setting. However, the strength of the observed association between pregnancy loss and postpartum mortality makes it an ideal variable to consider for risk stratification purposes, particularly as 60% of maternal deaths in our setting are potentially preventable.^5^ Women might suffer complications from abortions or miscarriages, which place them at increased risk of postpartum mortality.^5^ Pre-existing poor health or poorly controlled comorbidity not only contributes to pregnancy loss, but also has negative implications for maternal survival during the postpartum period. This is confirmed by previous reports of maternal mortality in South Africa, wherein non-pregnancy-related infections and hypertensive disease were identified as some of the most important causes of preventable maternal mortality.^5^ The psychosocial impact of pregnancy loss on mothers must also be considered, as published evidence links this adverse pregnancy outcome with subsequent maternal mental illness, self-destructive behavior, and suicide.^9, 14, 15^ Therefore, women who are identified as being at risk of postpartum mortality based on a recent history of pregnancy loss should be targeted with appropriate interventions which seek to timeously and efficiently manage medical complications from abortions or miscarriages, ensure adequate control of existing comorbid disease, and address the possible mental health consequences of pregnancy loss. These interventions might include regular, stringent clinical monitoring or follow-up of women who have had abortions or miscarriages; increasing access to care in women with pre-existing comorbidity, possibly through decentralized care in the community; and support groups for women struggling with the psychosocial consequences of pregnancy loss.^5, 16^

Our analysis also found that increasing age and an HIV-positive serostatus were associated with increased postpartum mortality, while a prior maternal history was associated with decreased postpartum mortality. Increasing age is a common predictor of maternal mortality throughout the world.^17^ Although our study found a significant increase in postpartum mortality amongst women in the 26-35 year age group, we were only able to observe a statistical trend toward increased postpartum mortality in women aged >35 years old. It is likely that there would also have been a statistically significant increase in postpartum mortality in women aged >35 years old had the sample of women in this specific age group been larger (only 6.2% of our sample was comprised of postpartum women >35 years old). Our finding for HIV serostatus confirms the importance of non-pregnancy-related infections as a cause of maternal mortality in South Africa.^5^ This finding is somewhat concerning, if one considers the high HIV prevalence amongst pregnant women in our setting and the challenge of ensuring that HIV infection in mothers is adequately controlled.^18^ The published literature suggests a “J-shaped” relationship between parity and maternal mortality, with nulliparous women and women with >7 prior pregnancies at highest risk for this outcome.^19^ Our study confirms this widely accepted relationship between nulliparity and increased maternal mortality. Thus, increasing age, HIV infection, and nulliparity are additional markers of increased postpartum mortality risk in our setting, which need to be considered during the risk stratification process.

The main strength of our study was that it was population-based and does not suffer from many of the biases typically associated with facility-based studies. This renders our findings more generalizable to pregnant women in sub-Saharan African settings. Another important strength of our study was that the data collection involved a “full pregnancy history” (FPH) questionnaire (which collects information on all pregnancies during a woman’s life, irrespective if these ended in live births or not) rather than the “full birth history” (FBH) questionnaire (which has traditionally collected information only on live births, with updates of the instrument in 2013 allowing for limited information on pregnancy loss to be collected).^20^ As such, the pregnancy outcomes data in this study is far more robust than if the data had been collected using a FBH questionnaire. The large study sample size of 25048 postpartum women, with complete follow-up data, also facilitated an appropriate statistical analysis. This was particularly important considering that only 3.8% of our study sample experienced pregnancy loss.

The main limitation of our study is the potential for selection bias among reported pregnancy outcomes. Information on pregnancy outcomes was self-reported by women who participated in the AHRI general household surveys, and the information regarding pregnancy losses is sensitive and potentially vulnerable to omission. ^21, 22^ As a result, it is possible that pregnancy losses are underreported during the AHRI general household surveys. While is difficult to measure how this affects our results, we hypothesize that the association between pregnancy loss and postpartum mortality would be even stronger in the absence of selection. This proposition is supported by the previously established relationship between women who experienced pregnancy loss and postpartum mortality, as well as the shared risk factors for such events.

In conclusion, our research from a typical rural sub-Saharan African setting demonstrates a three-fold higher incidence of postpartum mortality in women who had recently experienced pregnancy loss, when compared with women who had live births. Our results indicate that women who have recently experienced a pregnancy loss should be targeted with a package of risk reduction interventions, including opportunities for medical care or psychosocial support at regular intervals in the year following the end of their pregnancy.

## Supporting information

STROBE Checklist

## Data Availability

All datasets used in this study are publicly accessible through the Africa Health Research Institute's Data Repository.

https://data.ahri.org/index.php/home

## Acknowledgements

Yoshan Moodley contributed to the study’s conception, design, statistical analysis, and drafting of the manuscript. Hallie Eilerts reviewed the draft manuscript and provided expertise on the strengths and limitations of the AHRI pregnancy data used in this study. Kobus Herbst contributed to the data collection and drafting of the manuscript. Frank Tanser contributed to the study’s conception and drafting of the manuscript. All authors read and approved the final version of this manuscript. The authors are g rateful to the study participants and the work and support of the fieldwork and database teams at AHRI. The content of this manuscript is solely the responsibility of the authors and does not necessarily represent the official views of the funding bodies.

### Authors’ Data Sharing Statement

- Will individual participant data be available (including data dictionaries)? *Yes. All datasets used in this study are available from the AHRI Data Repository at* [https://data.ahri.org/index.php/home]: *Pregnancy dataset* [https://doi.org/10.23664/AHRI.RD01-04.COREDATASET.PREGNANCIES.201909], General Household Survey dataset [https://doi.org/10.23664/AHRI.RD07-99.PIP.HSE-I.ALL.201907], *HIV Surveillance dataset* [https://doi.org/10.23664/AHRI.RD07-99.PIP.HIV.ALL.202007].
- What data in particular will be shared? *The datasets on the AHRI data repository are de-identified and comprise individual-level information on various demographic and health variables*.
- What other documents will be available? *Brief overall description of the specific study/survey from which the data was obtained, as well as data dictionaries*.
- When will data be available (start and end dates)? *The datasets are currently available, pending any future changes in data access policies at AHRI*.
- By what access criteria will data be shared (including with whom, for what types of analyses, and by what mechanism)? *The datasets used in this study are available upon reasonable request from the AHRI Data Repository at* [https://data.ahri.org/index.php/home].

## APPENDIX 1

Flow diagram showing derivation of the study sample:

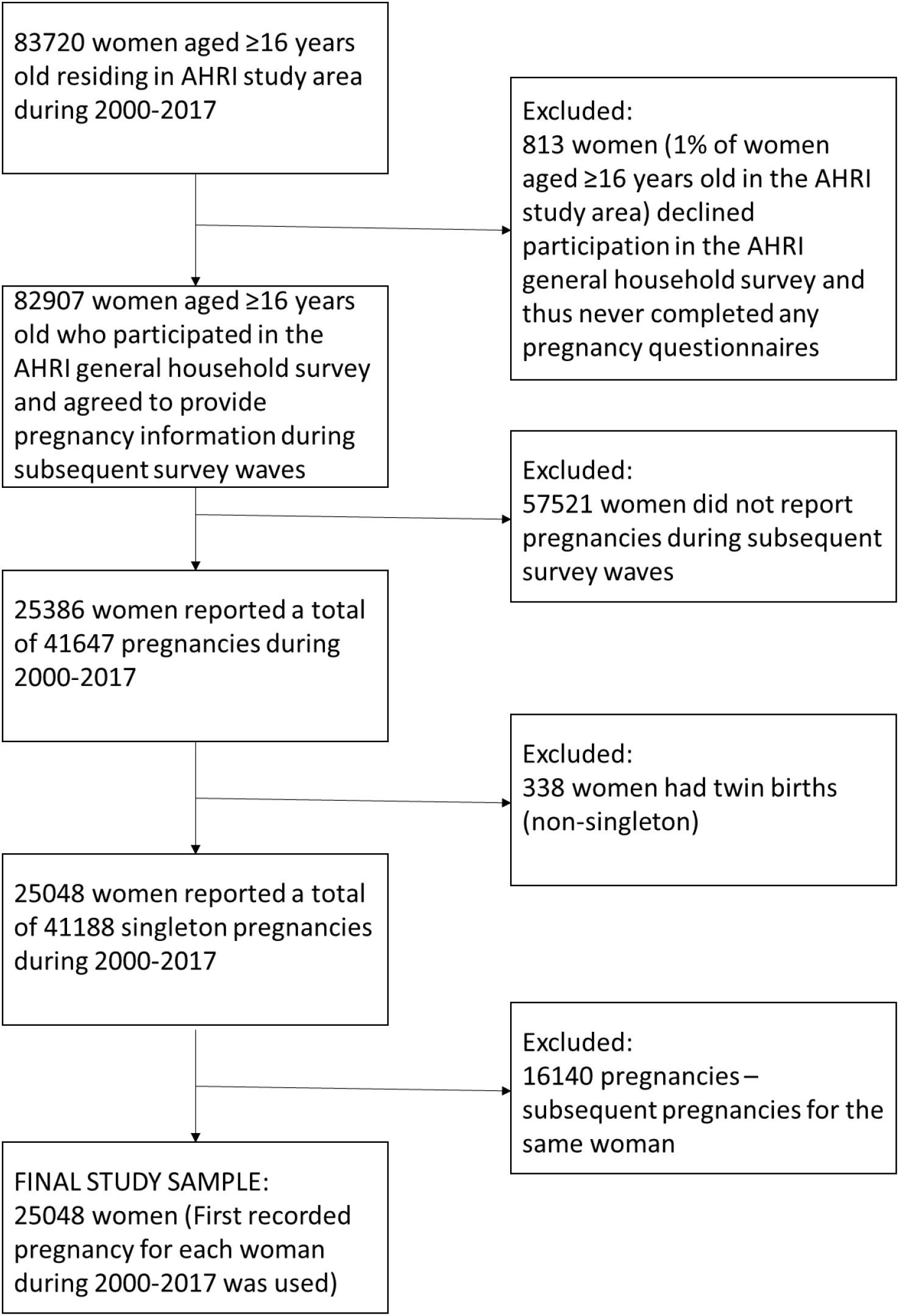

